# Altered transcriptome-proteome coupling indicates aberrant proteostasis in Parkinson’s disease

**DOI:** 10.1101/2021.03.18.21253875

**Authors:** Fiona Dick, Ole-Bjørn Tysnes, Guido Werner Alves, Gonzalo S. Nido, Charalampos Tzoulis

## Abstract

The correlation between mRNA and protein levels has been shown to decline in the ageing brain, possibly reflecting age-dependent changes in the proteostasis. It is thought that impaired proteostasis may be implicated in the pathogenesis of Parkinson’s disease (PD), but evidence derived from the patient brain is currently limited. Here, we hypothesized that if impaired proteostasis occurs in PD, this should be reflected in the form of altered correlation between transcriptome and proteome compared to healthy ageing.

To test this hypothesis, we integrated transcriptomic data with proteomics from prefrontal cortex tissue of 17 PD patients and 11 demographically matched healthy controls and assessed gene-specific correlations between RNA and protein level. To control for the effects of ageing, brain samples from 4 infants were included in the analyses.

In the healthy aged brain, we observed a genome-wide decreased correlation between mRNA and protein levels. Moreover, a group of genes encoding synaptic vesicle proteins exhibited inverse correlations. This phenomenon likely reflects the spatial separation of mRNA and protein into the neuronal soma and synapsis, respectively, commonly characterizing these genes. Most genes showed a significantly lower correlation between mRNA and protein levels in PD compared to neurologically healthy ageing, consistent with a proteome-wide decline in proteostasis. Genes showing an inverse correlation in PD were enriched for proteasome subunits, suggesting that these proteins show accentuated spatial separation of transcript and protein between the soma and axon/synapses in PD neurons. Moreover, the PD brain was characterized by increased positive mRNA-protein correlation for some genes encoding components of the mitochondrial respiratory chain, suggesting these may require tighter regulation in the face of mitochondrial pathology characterizing the PD brain.

Our results are highly consistent with a proteome-wide impairment of proteostasis in the PD brain and strongly support the hypothesis that aberrant proteasomal function is implicated in the pathogenesis of PD. Moreover, our findings have important implications for the correct interpretation of differential gene expression studies in PD. In the presence of disease-specific altered coupling of transcriptome and proteome, measured differences in mRNA levels cannot be used to infer changes at the protein-level and should be supplemented with direct determination of proteins nominated by the analyses.

## Introduction

Gene expression is the process by which the information encoded in the genome (i.e., the genotype) determines the phenotype. Typically, information encoded in the DNA is first transcribed to RNA and then translated into protein, the functional product influencing the phenotype [42]. Despite the hierarchical organization of gene expression, the relationship between transcript and protein levels is highly variable in mammalian cells, both across genes and across individuals. Imperfect correlations between mRNA and protein levels are commonly attributed to regulatory mechanisms acting downstream of transcription and influencing the rate of protein synthesis and degradation [5, 10, 28]. For example, splicing, polyadenylation, and RNA-binding factors regulate translation rates, while the ubiquitin-proteasome system and lysosomal degradation regulate protein turnover. The balanced interplay between these regulatory mechanisms is crucial for maintaining cellular proteostasis.

It was recently shown that the correlation between mRNA and protein levels declines with ageing in the human brain, possibly due to altered post-transcriptional regulation [8, 54] and declining proteostasis [28]. Impaired proteostasis is thought to contribute to the misfolding and aggregation of proteins observed in neurons and other postmitotic cells with ageing [28], a phenomenon that is substantially more pronounced in age-dependent neurodegenerative proteinopathies, such as Parkinson’s disease (PD) and Alzheimer’s disease (AD) [17]. Several lines of evidence indicate that aberrant proteostasis is indeed implicated in PD [36]. The accumulation of Lewy bodies and neurites, intraneuronal inclusions containing aggregated forms of the protein α-synuclein [27], suggests decreased function of the autophagy–lysosomal pathway [37]. This is further supported by the fact that mutations in *GBA*, encoding the lysosomal enzyme glucocerebrosidase, greatly increase the risk of PD [1]. Altered mRNA levels of proteasomal components have been consistently found in transcriptomic studies of the PD brain [7], suggesting that dysfunction of the ubiquitin-proteasome system may also play a role.

We hypothesized that if impaired proteostasis occurs in PD, this should be reflected in the form of altered correlation between the transcriptome and proteome in the patients’ brain compared to healthy ageing. To test our hypothesis, we performed transcriptome and proteome-wide analyses, using RNA sequencing and proteomics, in the brain of 17 PD patients and 11 demographically matched healthy controls, and assessed the correlation between the levels of each transcript and its cognate protein. Since it is known that extensive changes leading to mRNA-protein decoupling occur with ageing in the human brain [8, 54], we also analyzed brain samples of four individuals in early infancy. Ageing remains the strongest known PD risk factor, and this additional group allowed us to distinguish changes in mRNA-protein correlations arising due to neurologically healthy ageing from those that are specific to pathological ageing with PD.

Our results show that the PD brain is characterized by genome-wide altered mRNA-protein correlation, compared to neurologically healthy ageing. The pattern of this altered relationship between transcriptome and proteome is highly consistent with a disease-related impairment in proteostasis.

## Materials and Methods

### Cohorts

All experiments were conducted in fresh-frozen prefrontal cortex (Brodmann area 9) tissue from a total of 33 individuals comprising young infants (YG, *N* = 4, age 0-0.38 years), neurologically healthy aged individuals (HA, *N* = 11, age 63-88 years) and individuals with idiopathic Parkinson’s disease (PD) (*N* = 17, age 69-95 years) from the Park-West study, a prospective population-based cohort which has been described in detail [2]. Whole-exome sequencing had been performed on all PD patients and known causes of Mendelian PD and other monogenic neurological disorders had been excluded [19]. Controls had no known neurological disease and were matched for age and gender. Individuals with PD fulfilled the National Institute of Neurological Disorders and htroke [20] and the UK Parkinson’s disease Society Brain Bank [53] diagnostic criteria for the disease at their final visit. Ethical permission for these studies was obtained from our regional ethics committee (REK 2017/2082, 2010/1700, 131.04). Cohort demographics are listed in S1 File.

### Sample collection

Brains were collected at autopsy and split sagittally along the corpus callosum. One hemisphere was fixed whole in formaldehyde and the other coronally sectioned and snap-frozen in liquid nitrogen. All samples were collected using a standard technique and fixation time of ∼ 2 weeks. Subject demographics and tissue availability are provided in S1 Figure. Routine neuropathological examination including immunohistochemistry for *α*-synuclein, tau and beta-amyloid was performed on PD and HA brains. All PD cases showed neuropathological changes consistent with PD including degeneration of the dopaminergic neurons of the substantia nigra pars compacta in the presence of Lewy pathology. Controls had no pathological evidence of neurodegeneration.

### RNA sequencing

Total RNA was extracted from prefrontal cortex tissue homogenate for all samples using RNeasy plus mini kit (Qiagen) with on-column DNase treatment according to the manufacturer’s protocol. The final elution was made in 65*µ*l of dH2O. The concentration and integrity of the total RNA were estimated by Ribogreen assay (Thermo Fisher Scientific), and Fragment Analyzer (Advanced Analytical), respectively and 500ng of total RNA was used for downstream RNA-seq applications. First, nuclear and mitochondrial rRNA was removed using Ribo-Zero™ Gold (Epidemiology) kit (Illumina, San Diego, CA) using the manufacturer’s recommended protocol. Immediately after rRNA removal, RNA was fragmented and primed for the first strand synthesis using the NEBNext First Strand synthesis module (New England BioLabs Inc., Ipswich, MA). Directional second strand synthesis was performed using NEBNExt Ultra Directional second strand synthesis kit. Following this, the samples were taken into standard library preparation protocol using NEBNext DNA Library Prep Master Mix Set for Illumina with slight modifications. Briefly, end-repair was done followed by poly(A) addition and custom adapter ligation. Post-ligated materials were individually barcoded with unique in-house Genomic Services Lab (GSL) primers and amplified through 12cycles of PCR. Library quantity was assessed by Picogreen Assay (Thermo Fisher Scientific), and the library quality was estimated by utilizing a DNA High Sense chip on a Caliper Gx (Perkin Elmer). Accurate quantification of the final libraries for sequencing applications was determined using the qPCR-based KAPA Biosystems Library Quantification kit (Kapa Biosystems, Inc.). Each library was diluted to a final concentration of 12.5nM and pooled equimolar prior to clustering. One hundred twenty-five bp Paired-End (PE) sequencing was performed on an Illumina HiSeq2500 sequencer (Illumina, Inc.). RNA quality, measured by the DV200 score, varied across samples (*median*_*Y G*_ = 92, *median*_*CT*_ = 88, *median*_*P D*_ = 89), although the difference between groups was not statistically significant (*p*_*Y G,CT*_ = 0.06, *p*_*CT,P D*_ = 0.74, *p*_*Y G,P D*_ = 0.07, Wilcoxon rank sum test).

### RNA-Seq quality control and transcript abundance estimation

FASTQ files were trimmed using Trimmomatic version 0.39 [6] to remove potential Illumina adapters and low quality bases with the following parameters: `ILLUMINACLIP:truseq.fa:2:30:10 LEADING:3 TRAILING:3 SLIDINGWINDOW:4:15`. FASTQ files were assessed using fastQC version 0.11.5 [3] prior to and following trimming. We used Salmon version 1.3.0 [41] to quantify the abundance at the transcript level with the fragment-level GC bias correction option (--gcBias) using the GENCODE Release 32 (GRCh38.p13) reference transcriptome and the GRCh38 reference genome, included as decoy [47]. Transcript counts were collapsed to gene-level using R package tximport version 1.14.2 with default parameters (i.e., countsFromAbundances = FALSE) and the GENCODE Release 32 (GRCh38.p13) annotation. Henceforth, we use the notion of *transcript* in a gene-centric sense, i.e., as the entity defined by all transcript isoforms mapped to the same gene. mtDNA-encoded genes were removed from the analysis. Genes were further filtered out if unusually highly expressed (i.e., if they accounted for more than 1% of a sample’s library size in more than 50% of all the neurologically healthy samples (i.e., YG, HA)). We calculated *log*_2_ transformed counts per million (CPM) for the pre-filtered set of genes. Low-expressed genes (*log*_2_ − *CPM* < 0.1, in at least 80% of the samples) were also filtered out. The pre-filtered transcriptomic dataset resulted in a total of *N* = 29, 601 genes. The dataset corresponding to the PD samples, added subsequently in the analyses, was filtered independently following the same filtering approaches and resulting in a total of *N* = 29, 363 genes.

### Lysis and protein digestion

10*µL* of lysis buffer (4% SDS, 0.01*M* TRIS pH 7.6) was added to 1*mg* of brain tissue. The tissue was mechanically lysed using Precellys CK 14 ceramic beads, together with the Precellys Evolution (Bertin Corp, Rockville MD, USA). Lysed tissue was transferred to Eppendorf tubes and heated to 95°C for 5 minutes, before centrifugation at 10.000*g* for 5 minutes. The clarified supernatant was transferred to new Eppendorf tubes. Protein measurement was performed using the Pierce BCA protein assay kit (Thermo Fisher).

The samples were mixed with up to 50*µL* of the clarified lysate with 200*µL* of 8 M urea in 0.1 M Tris/HCl pH 8.5 in the filter unit (Microcon YM-30 (Millipore, Cat. MRCF0R030)) and centrifuged at 14,000 × g for 30 min and repeated twice. In total 30*µg* of protein per sample was used. The samples were reduced with 10*mM* DTT (1h, RT) and alkylated using 50*mM* IAA (1h, RT), and digested overnight at 37°C with 1:50 enzyme: substrate ratio of sequencing grade trypsin (Promega, Madison, WI). Following digestion, samples were acidified with formic acid and desalted using HLB Oasis SPE cartridges (Waters, Milford, MA). Samples were eluted with 80% acetonitrile in 0.1% formic acid and lyophilized. Peptides were stored at −80°C until use [26].

### TMT labeling and fractionation

Digested peptides from each sample were chemically labelled with TMT reagents 10 plex (Thermo Fisher). Peptides were resuspended in a 30*µL* resuspension buffer containing 0.1*M* TEAB (Triethylammonium bicarbonate). TMT reagents (0.1*mg*) were dissolved in 41*µL* of anhydrous ACN of which 20*µL* was added to the peptides. Following incubation at RT for 1 h, the reaction was quenched using 5% hydroxylamine in HEPES buffer for 15 min at RT. The TMT-labeled samples were pooled at equal protein ratios followed by vacuum centrifuge to near dryness and desalting using Oasis PRIME HLB cartridges. Peptides were fractionated into 8 fractions using the Pierce High pH Reverse-phase Peptide fractionation kit (Thermo Fisher Scientific). The TMT experiment batch setup included additional samples which were not considered in the analysis but included in the preprocessing (filtering and normalization) of the proteomics data.

### Liquid Chromatography and Mass Spectrometry Analysis

Each sample was freeze-dried in a Centrivap Concentrator (Labconco) and dissolved in 2% ACN, 1% FA. Approximately 0.5 *µ*g of peptides from each fraction was injected into an Ultimate 3000 RSLC system (Thermo Scientific) connected to a Q-Exactive HF equipped with an EASY-spray ion source (Thermo Scientific). The samples were loaded and desalted on a precolumn (Acclaim PepMap 100, 2 cm ·75*µm* i.d. nanoViper column, packed with 3 *µ*m C18 beads) at a flow rate of 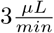 for 5 min with 0.1% TFA. The peptides were separated during a biphasic ACN gradient from two nanoflow UPLC pumps (flow rate of 0.200 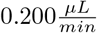) on a 50*cm* analytical column (PepMap RSLC, 50 cm ·

75*µm* i.d. EASY-spray column, packed with 2*µm* C18 beads (Thermo Scientific). Solvent A was 0.1% FA in water, and Solvent B was 100% ACN. The mass spectrometer was operated in data-dependent acquisition mode to automatically switch between full scan MS1 and MS2 acquisition. The instrument was controlled through Q Excative HF Tune 2.4 and Xcalibur 3.0. MS spectra were acquired in the scan range of 375 − 1500 m/z with resolution of 60, 000 at m/z 200, automatic gain control (AGC) target of 3 · 10^6^, and a maximum injection time (IT) of 50*ms*. The 12 most intense eluting peptides above intensity threshold 6 · 10^4^, and charge states two or higher, were sequentially isolated for higher energy collision dissociation (HCD) fragmentation and MS2 acquisition to a normalized HCD collision energy of 32%, target AGC value of 1 10^5^, resolution *R* = 60, 000, and IT of 110 ms. The precursor isolation window was set to 1.6*m/z* with an isolation offset of 0.3 and a dynamic exclusion of 30*s*. Lock-mass (445.12003 m/z) internal calibration was used, and isotope exclusion was active.

Raw data were analyzed by MaxQuant v1.5.5.1 [13] with “Variable Modifications” set for TMT 10-plex 126, 127N, 127C, 128N 128C, 129N, 129C, 130N, 130C, 131 to be at N-termini, as well as lysine for database searching and peptide identification.

### Proteomics normalization and filtering

Aggregated protein intensities from maxQuant were further processed in a downstream analysis using R. First, proteins labelled as “Reverse”, “Potential. contaminant” and “Only.identified.by.site” were removed from the analysis. In addition, proteins were removed if they exhibited at least one zero intensity in a sample. In order to filter out highly-expressed proteins, we selected the top four highest expressed proteins in each sample (which ranged from 3% to 5% of the total expression of a sample). The union set of these (a total of 19 proteins) was then filtered out from every sample.

We considered three possible normalization approaches for protein quantification, i) raw protein intensities, ii) quantile normalization, and iii) batch effect correction [9] followed by root mean square scaling. To assess each of these strategies we explored the association of the first two components of the principal component analysis (PCA) of the protein expression matrix with the TMT batch. Raw protein intensities (i) showed a clear clustering of samples which was associated with the batches of the TMT experiment, which was further amplified by quantile normalization (ii). This effect was no longer noticeable when we applied batch correction (iii), as suggested in [9], where we divided protein intensities by the correction factor based on the reference channels in the respective batches, followed by root mean square scaling (S1 Figure). Additionally, we were able to leverage the RNA-seq data from the same samples to gain insight into the biological validity of the three alternative normalization options by studying the transcriptome-proteome correlation in the neurologically healthy groups (HA and YG; *log*_2_ transformed values for proteins, and *log*_2_ transcript CPMs). The transcriptome-proteome correlation was significantly higher in the batch-corrected strategy both across samples and across genes (S2 Figure). Based on these observations we chose to apply the batch correction and subsequent root mean square scaling (iii). The pre-filtered proteomic dataset was composed of a total of *N* = 2, 961 proteins.

### Covariance between omic layers

We used sparse partial least square (sPLS) as implemented in the mixOmics R package version 6.10.9 [34, 43] to find the linear combinations of variables (transcripts and proteins) that maximize covariance between the transcriptomic and the proteomic layers. sPLS was performed on the pre-filtered transcriptomic (X) and proteomic (Y) datasets using the “canonical” mode and the parameters keepX = 50 and keepY = 50 for feature selection.

### Correlation between transcriptome and proteome

To investigate changes in the transcriptome-proteome correlation between neurologically-healthy groups (YG *vs* HA) we performed an additional filtering step on both transcripts and proteins, aiming at increasing the biological signal-to-noise ratio. Genes were flagged for removal if they satisfied at least one of the following criteria: i) not present in the pre-filtered transcriptome, ii) not present in the pre-filtered proteome, iii) low median transcript expression (below 10% quantile), iv) low transcript variance (below 15% quantile). The removal of flagged genes resulted in an analysis-ready dataset of *N* = 2, 107 genes.

The dataset corresponding to the PD samples, employed in a subsequent comparison, was filtered independently following the same filtering approaches and resulting in a slightly lower number of genes in the final analysis-ready list (*N* = 1, 942). Gene-wise transcript-protein Pearson correlations were calculated across samples independently for each group (CT, PD, YG) using *log*_2_ transformed CPMs for transcript abundance and log2 transformed batch-corrected and root mean square scaled protein intensities.

### Gene scoring

To investigate changes in the transcriptome-proteome correlation between groups, we applied different gene scoring strategies to rank genes according to their change in correlation (*δr*). For example, to investigate changes occurring in the healthy ageing process (i.e., comparing YG vs HA) each gene would be scored by *δr* = *r*_*HA*_ − *r*_*Y G*_. Correspondingly, to investigate changes occurring in the process of ageing with Parkinson’s disease, gene scores would be calculated as *δr* = *r*_*PD*_ − *r*_*YG*_. Finally, changes in transcript-protein correlations between CT and PD groups would be calculated as *δr* = *r*_*P D*_ − *r*_*HA*_ (Figure 1A).

Specifically, for each of these three group comparisons (YG→HA, YG→PD, HA→PD), we wanted to identify genes belonging to three functional scenarios in regard to their transcript-protein coupling: a) *”decoupling”*, genes that show a positive transcript-protein correlation in the reference group (e.g., YG) and loose this correlation (*r ∼* 0) in the other group (e.g., HA); b) *“increased inverse correlation”*, genes which show a correlation above or equal to zero in the reference group and a negative correlation in the other group; and c) *“increased positive correlation”*, genes with a correlation above or equal to zero in the reference group that show an increased correlation in the group compared (Figure 1B). To this end, gene-specific scores were calculated as follows:

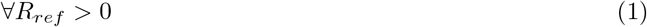

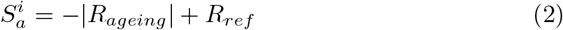

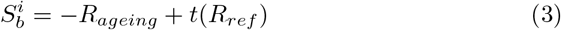

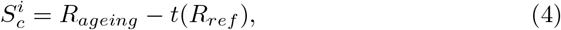

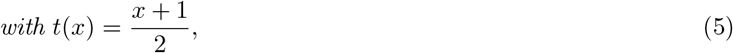

where *i* ∈ 1, 2, 3 specified the comparison being made:

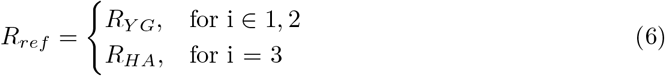

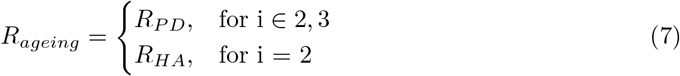

Heatmaps to visualize scoring distributions in Figure 1C were created with the R package ComplexHeatmap [23].

**Figure 1.**
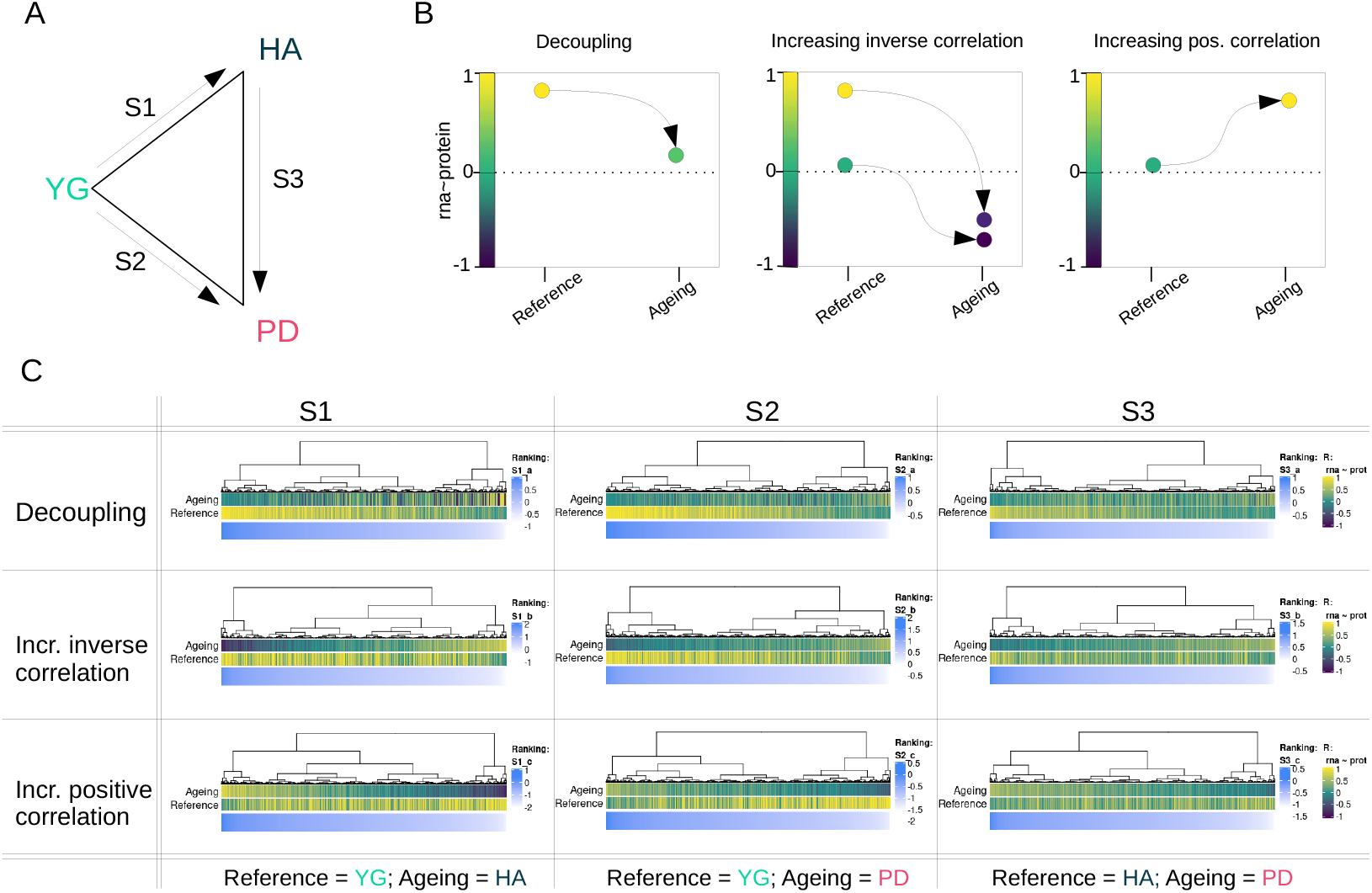
Gene scoring ranking for gene-set enrichment analysis. **A:** Schematic illustration of comparisons between groups. Each comparison is between a reference group and an ageing group (either HA or PD). For S1, we define YG as the reference and HA as the ageing group. Similarly, for S2, we define YG as a reference and PD as the ageing group. Finally, in S3 we investigate the differences between HA (reference) and PD (ageing). PD: Parkinson’s disease; HA: healthy aging; YG: infants. **B:** Schematic representation of correlation changes: i) decoupling ii) increasing inverse correlation and iii) increasing positive correlation. We calculated scores to rank genes according to each of these three trends to perform change-specific pathway enrichment analysis. **C:** Gene scores calculated for the three comparisons (as defined in A) and correlation trends (as defined in B) displayed in blue, mapped to the respective reference and ageing correlation coefficient. The correlation coefficients are coloured from −1 (dark blue) to zero (green) to 1 (yellow)

### Pathway enrichment analysis and clustering for visualization

The above gene scorings were used to test for functional enrichment. To this end, we employed the gene score resampling method implemented in the R package ermineR version 1.0.1.9, an R wrapper package for ermineJ [35] with the complete Gene Ontology (GO) database annotation [4] (using aspects: biological process, molecular function and cellular component).

### Protein interaction networks

Protein-protein interaction networks were generated using the R package coexnet version 1.8.0 [25], which retrieves information on protein co-expression and experimentally evidenced interaction from STRING [50]. Vertices were clustered using the R package igraph version 1.2.5 [14], and its implemented “edge-betweenness” cluster algorithm.

## Results

### Brain RNA and protein expression patterns are highly distinct between neurodevelopment and healthy ageing

Using RNA-seq and LC-MS-based proteomics, we mapped the transcriptome and proteome in prefrontal cortex tissue from 4 young infants (YG), 11 neurologically healthy aged individuals (HA), and 17 individuals with idiopathic PD (PD). First, we assessed the overall expression pattern of the groups YG and HA by integrating gene expression (X, *N* = 29, 601 genes) with protein expression (Y, *N* = 2, 918 proteins). Using sparse Partial Least Squares regression (sPLS), we were able to reduce dimensionality for both X and Y and project the samples in an unsupervised manner onto the combined XY-variate space. The groups YG and HA were markedly separated according to their biological characteristics in the combined variate space (YG cluster median silhouette width = 0.71, Euclidean distance; HA group median silhouette width = 0.53, Euclidean distance; Figure 2A) as well as in the separated variate space (Figure 2B), meaning that the group separation was independent of whether the selected features were restricted to either the transcriptome or the proteome, with both datasets strongly agreeing. The first XY-variate, was strongly correlated with age (*r* = 0.95, *p* = 8.24 · 10^*−*8^, Pearson). The *N* = 50 selected features for each component (I, II) of X and Y, which were sufficient to separate the groups, are visualized in a correlation heatmap in Figure 2C, highlighting interactions between features of X and features of Y for which the correlation was greater than *r* = 0.2.

**Figure 2.**
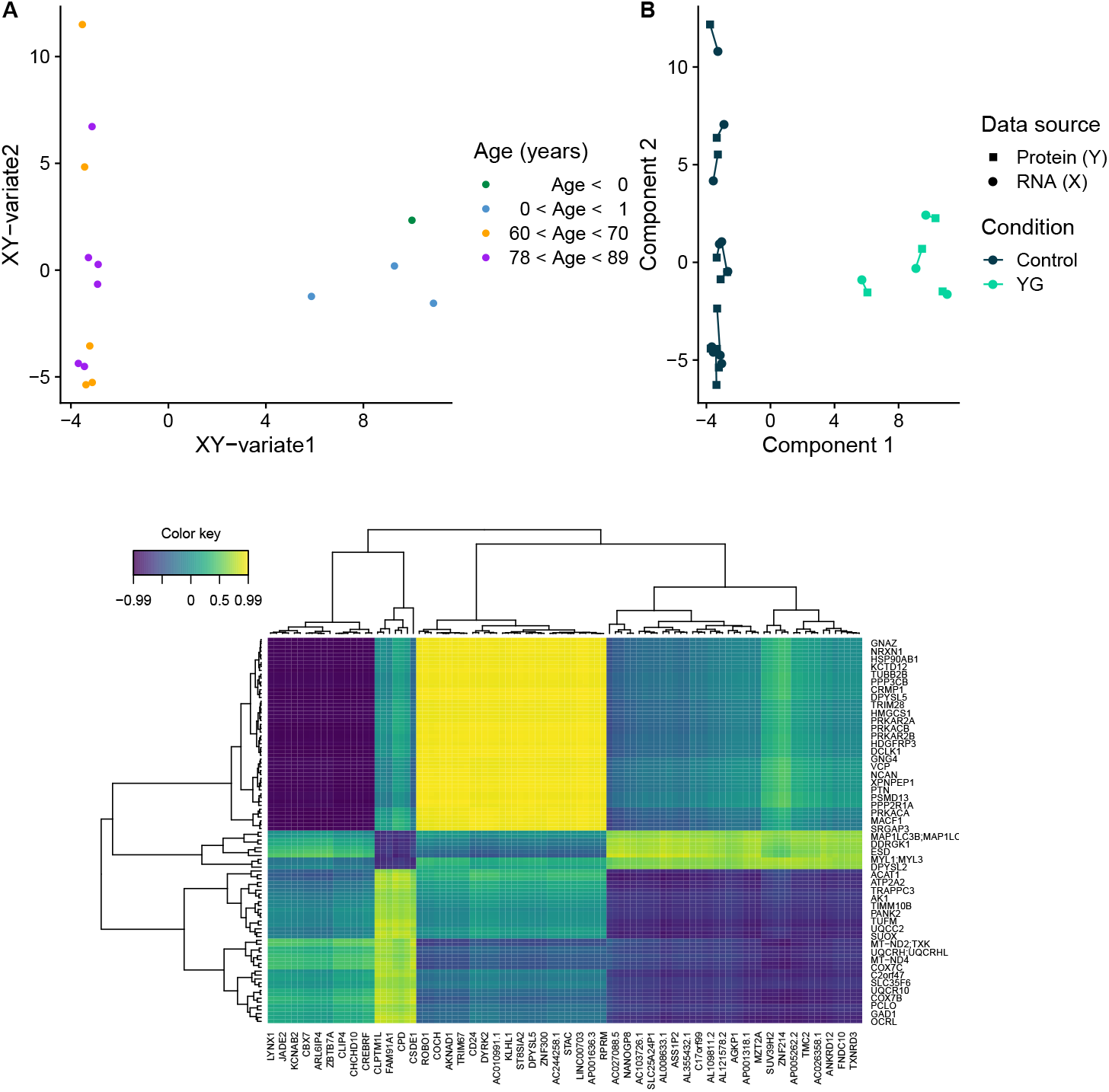
Integrative analysis of age-specific expression patterns in the transcriptome and proteome using sPLS. **A:** Data points (samples) coloured by age in years (binned) in the combined XY variate space (coordinates of samples are the mean over the coordinates in the subspaces of X and Y). **B:** Samples plotted separately in the subspaces X (circle) and Y (square) spanned by their first two components. Colour coding indicates group membership (HA: dark blue; YG: turquoise); shape indicates omic layer (protein expression: square; transcript expression: circle). **C:** Heatmap displaying the selected features of components I and II from both omic layers: transcriptome (x-axis) and proteome (y-axis). A correlation threshold of 0.2 was set to reduce the number of features in the plot and facilitate visualization. Colour indicates correlation between features of X and features of Y.

### The transcriptome-proteome correlation signature is altered in the aged brain

Since mRNA and protein levels are known to be tightly correlated during neurodevelopment, we leveraged the YG group as a control outgroup to assess alterations that occur with age and/or PD. To compare the transcriptome-proteome coupling between YG and HA groups, we calculated gene-wise correlation coefficients (*r*, Pearson) across samples in each of the groups (*r*_*Y G*_ and *r*_*HA*_) for the YG and HA groups, respectively). After pre-filtering, we were able to assess the transcript-protein level correlation for 2,107 genes. Correlation coefficients for each group are listed in S2 File. We will henceforth use the term *gene* for both the gene and the protein it encodes.

As expected, transcriptome and proteome were significantly more correlated in the YG group compared to the HA group as shown by the transcript-protein *r* distributions (median *r*_*Y G*_ = 0.31; median *r*_*HA*_ = 0.07; *p* < 2 · 10^*−*16^, Wilcoxon) (Figure 3A). To further characterize the differences in the transcriptome-proteome coupling, we generated a two-dimensional density plot of the gene-wise transcript-protein correlation in the YG and HA groups (Figure 3B). The vast majority of genes exhibited a high transcript-protein correlation in YG (*r*_*Y G*_ > 0.5) and a lack of correlation in HA (*r*_*HA*_$*sim*0). We henceforth refer to this age-dependent decrease in transcript-protein correlation as decreased coupling or, simply, *decoupling*. Additional high-density areas were observed for genes with low absolute transcript-protein correlation in both groups, and for genes transitioning from a highly positive correlation in YG to an inverse correlation in HA. Finally, very few genes showed an age-dependent increase in correlation. These observations indicate that most genes show a tight positive correlation between mRNA and protein levels during early infancy. With aging, however, this correlation either decreases towards zero (*r*_*HA*_ → 0, decoupling) or becomes inverse (*r*_*HA*_ < 0, increased anticorrelation).

**Figure 3.**
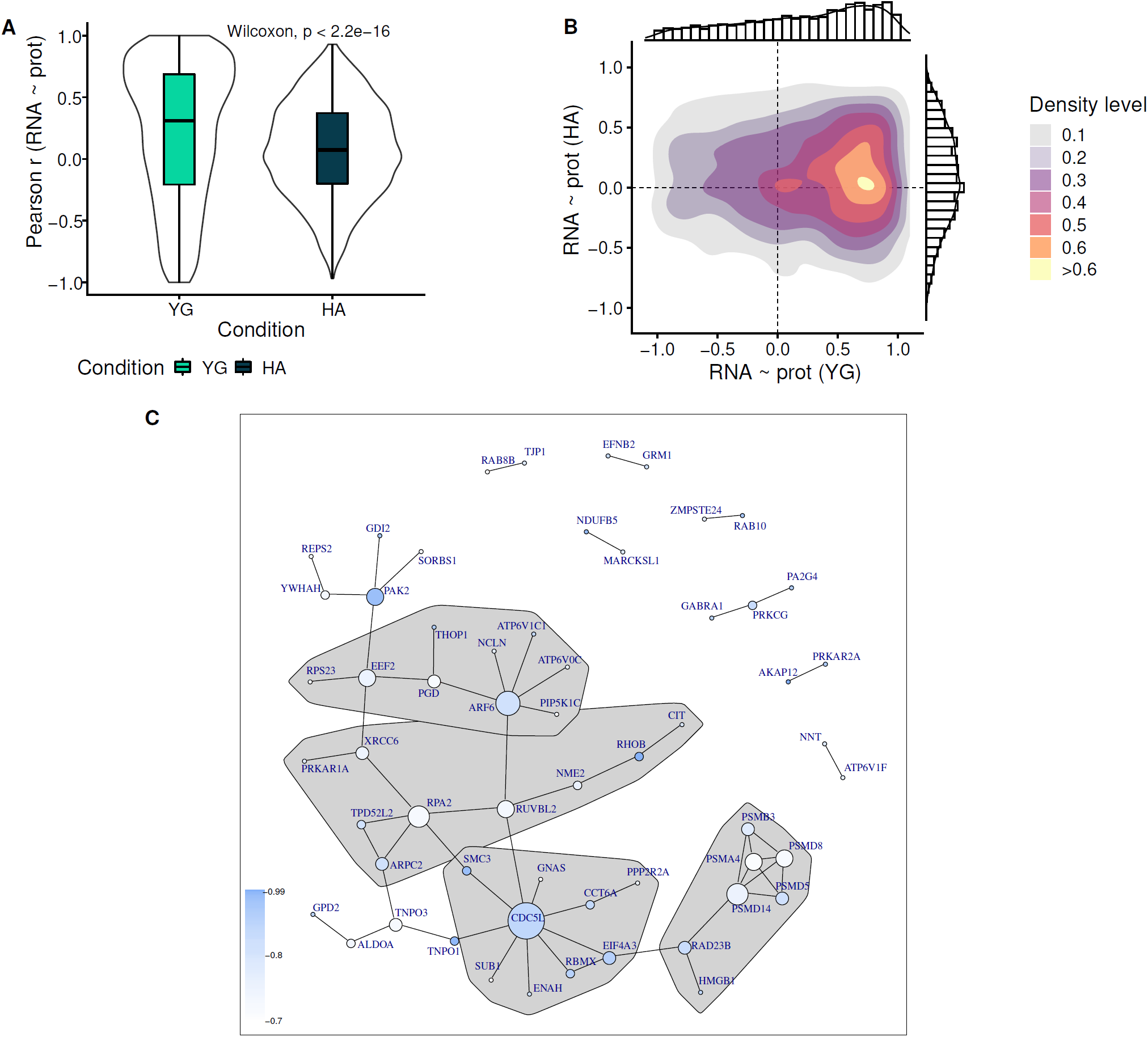
Decoupling of transcriptome and proteome in neurologically healthy aged individuals. **A:** Dist ribution of gene-wise correlation coefficients for the groups YG (turquoise) and HA (dark blue) (Wilcoxon unpaired test). **B:** Two-dimensional density plot displaying within-gene mRNA-protein Pearson correlations in YG (x-axis) vs HA (y-axis). **C:** Protein-protein interaction (PPI) network for genes in the 0.90 quantile of gene-scores ranking (blue) genes by decoupling in HA. Vertex communities were identified using edge betweenness (R package igraph). Only communities with more than 5 members are displayed. PPI based on coexpression, experimental evidence of interaction and neighbourhood characteristics.

### Altered mRNA-protein correlation in the aged brain is enriched in specific biological functions

Next, we assessed whether altered mRNA-protein correlation in the aged brain is enriched for specific pre-defined biological pathways. To this end, we divided genes into three groups according to their changes in correlation (YG → HA): i) decoupled (*r*_*Y G*_ > 0, *r*_*HA*_ ∼ 0), ii) increased inverse correlation (*r*_*Y G*_ > 0, *r*_*HA*_ < 0), iii) increased positive correlation (*r*_*Y G*_ > 0, *r*_*HA*_ > *r*_*Y G*_). Genes in each group were ranked according to the magnitude of the difference (*δ*(*r*_*HA*_, *r*_*Y G*_)) (Figure 1, S2 File). While the majority of genes showed decoupling with ageing (group i), we found no significant enrichment in this group for any specific biological pathway. A protein-protein interaction network of the top decoupled genes (gene score > 90% quantile, *N* = 58), revealed 4 interconnected groups with more than 5 members (Figure 3C), strongly suggesting a functional relationship. Notably, 5 of the 7 members of one of these groups were subunits of the proteasome complex (*PSMA4, PSMB3, PSMD5, PSMD8, PSMD14*). The gene group with increased inverse correlation (group ii) showed significant enrichment for 14 pathways (FDR < 0.05), mostly related to synaptic components and synaptic vesicles (S2 Figure, sheet S1b). Finally, the minority of genes which showed increased positive correlation from YG to HA (iii) were enriched in “regulation of hemostasis” (FDR = 0.04). Significantly enriched GO terms for each of the three groups are listed with their FDR adjusted p-value in (S2 Figure, sheet S1b-c).

### The age-dependent decoupling between mRNA and protein levels is more pronounced in the PD brain

Next, we wanted to assess how the coupling between transcriptome and proteome changes in PD compared to normal, neurologically healthy ageing. Transcript-protein correlations across all three groups (YG, HA and PD) were assessed for a total of 1,907 genes (see Methods). The correlation distributions for PD and HA groups showed no significant difference (*p* = 0.52, Wilcoxon) with a median close to zero for both groups (median *r*_*P D*_ = 0.10, median *r*_*HA*_ = 0.08). However, PD exhibited an overall lower variance (*σ*^2^(*r*_*HA*_) = 0.15, *σ*^2^(*r*_*P D*_) = 0.08) and a reduced range (*range*(*r*_*HA*_) = [–0.97, 0.93]; *range*(*r*_*P D*_) = [–0.70, 0.80]), suggesting a more pronounced trend of decoupling (Figure 4A).

**Figure 4.**
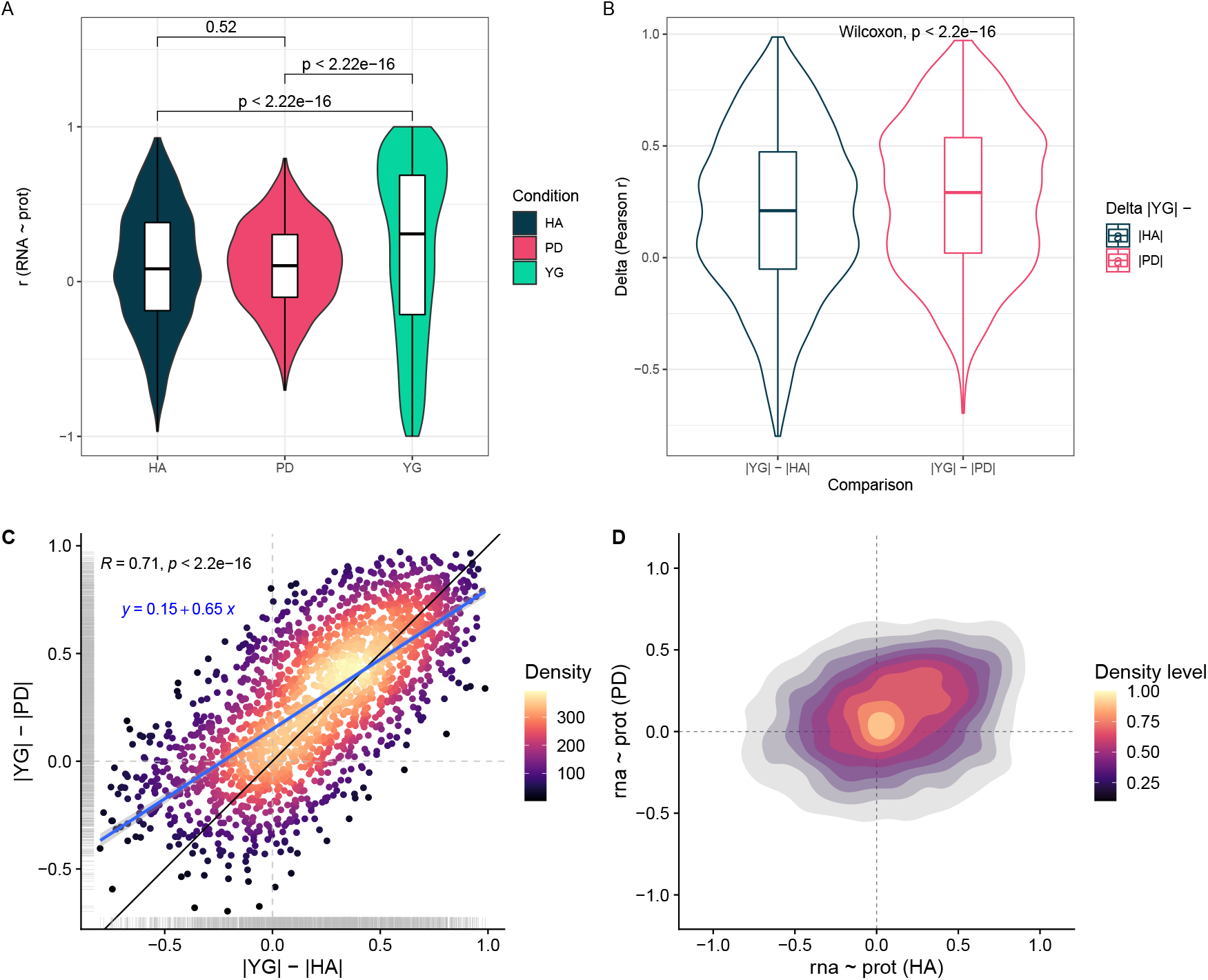
Altered correlation coefficient distribution in PD. **A:** mRNA-protein correlation distributions for HA (dark blue), PD (pink) and YG (turquoise). **B:** Distribution of the deltas (differences in absolute correlation coefficients) between the reference (YG) and HA (dark blue), and YG and PD (pink). **C:** Relationship between *δ*_*age*_ (x-axis) and *δ*_*P D*_ (y-axis). Color indicates data point density. Blue line indicated the linear model fit (*y* ∼ *x*). Black line is the diagonal (intercept = 0, slope = 1) **D:** Two-dimensional density plot displaying both distribution and relationship between the RNA ∼ protein correlations in HA (x-axis) and PD (y-axis).

To further investigate this, we calculated the absolute difference in the gene-wise transcript-protein correlation between the YG group and either the HA (*δ*_*age*_ = |*r*_*HA*_| − |*r*_*Y G*_|) or the PD group (*δ*_*P D*_ = |*r*_*P D*_| − |*r*_*Y G*_|). Interestingly, the two distributions differed significantly (*p* = 2.2 ·10^*−*16^, Wilcoxon, paired), with *δ*_*P D*_ being significantly higher than *delta*_*age*_ (Figure 4B). These findings indicate that the age-dependent loss of transcript-protein correlation is more pronounced in pathological aging with PD than in healthy aging, as evident also by the *r*_*HA*_*∼ r*_*Y G*_ and *r*_*P D*_*∼ r*_*Y G*_ density distributions (S3 Figure). Despite these differences, *δ*_*age*_ and *δ*_*P D*_ showed a highly significant positive correlation (*r* = 0.71, *p* < 2.2· 10^*−*16^, Pearson) (Figure 4C), suggesting that the process of decoupling is qualitative similar and has a comparable genome-wide distribution in HA and PD, although it is more pronounced in the latter.

### Altered transcript-protein correlation in the PD brain is enriched for specific biological processes

Similar to healthy ageing, transcript-protein correlation in the PD-brain showed a general trend for decoupling compared to the YG group (Figure 4D) and no significant enrichment for specific pathways. Genes showing increased inverse correlation with PD ageing were significantly enriched for 136 pathways (FDR < 0.05) mainly related to protein degradation (including proteasome complex, ubiquitination and unfolded protein response), immune response, transcription and cell-cycle regulation (S2 Figure, sheet S2b). However, a closer look at the enrichment results singled out several proteasomal subunits as main drivers of the enrichment signal across all these pathways. Removal of proteasomal subunits from the gene lists, in fact, resulted in only three statistically significant pathways. The signal of two of them (related to enhancer-binding), was driven solely by 5 genes (*CALCOCO1, H3F3A, RUVBL2, SFPQ, SUB1*), while the enrichment of the third significant pathway (unfolded protein binding) was supported by 36 genes which showed negative correlation in PD. Genes showing increased positive correlation with PD ageing were significantly enriched for 46 pathways, most of which were related to mitochondrial respiration (S2 Figure, sheet S2c).

Since comparing YG and PD cannot confidently differentiate ageing-from disease-related changes, we also performed a direct comparison between HA and PD. These analyses revealed an altered profile of transcript-protein correlation in PD compared to HA (Figure 4D). Genes showing increased decoupling in PD were significantly enriched for 17 pathways mostly related to nitrogen metabolism. Genes showing increased inverse correlation in PD were enriched in 54 biological pathways, primarily related to protein degradation and immune response. Similar to the comparison with the YG-group, this enrichment was driven primarily by proteasomal subunits. The magnitude of anticorrelation was, however, heterogeneous, affecting certain subunits more than others (*median*(*r*) = —0.18, *range*(*r*) = [− 0.64, 0.42], *σ*_*r*_ = 0.27, *N* = 22). Finally, genes with increased positive correlation in PD were enriched for biological processes related to mitochondrial respiration. A list of significantly enriched pathways in PD compared to HA is provided in S2 Figure, sheet S3a-c.

## Discussion

Here, we assess for the first time the genome-wide correlation between the transcriptome and proteome in the PD brain, compared to neurologically healthy ageing. In the infants, the vast majority of genes showed a strong positive correlation between mRNA and protein levels, suggesting that in the neonatal brain, protein abundance is determined mainly by transcript concentration. This correlation was significantly lower in the neurologically healthy aged individuals, consistent with an age-dependent decoupling between transcript and protein abundance. Similar trends have been shown in yeast [31], fish [32], and the macaque and human brain [8, 54]. Previous studies have suggested that age-dependent decoupling in the brain may preferentially affect certain biological processes, including transcriptional, translational and posttranslational regulation, signalling pathways, and mitochondrial function [31, 32, 54]. In our data, genes that decoupled in the aged group did not exhibit a significant enrichment in any specific biological pathways, suggesting that the age-dependent loss of correlation between mRNA and protein is a general, genome-wide process.

The phenomenon of age-dependent decoupling between mRNA and protein suggests that, in the ageing brain, modulating the rates of translation and protein degradation assumes a more central role in determining protein abundance than transcriptional regulation. On the other hand, the observed tight correlation between mRNA and protein levels in the neonatal brain may be, at least partly, related to the ongoing proliferation and migration of glial progenitors [49], a process heavily dependent on transcriptional regulation via the binding of a broad spectrum of transcription factors [15].

In addition to the physiological effects of brain development, the mRNA-protein decoupling observed in the ageing brain may also reflect pathological changes taking place in ageing postmitotic cells. A decline in proteasome function with ageing has been shown in multiple mammalian tissues and is believed to be contributing to the accumulation of misfolded and damaged proteins in the ageing brain (reviewed in [51]). Although not statistically significant, several subunits of the proteasomal complex were among the top decoupled during healthy ageing. Our findings provide further support to the notion of aberrant proteasomal function in the aged brain.

The age-dependent decoupling between mRNA and protein levels was significantly more pronounced in the brain of individuals with PD. While our data cannot elucidate the molecular mechanisms underlying this phenomenon, a state of heightened decoupling is consistent with disease-related impairment in proteostasis due to altered proteasomal and/or lysosomal function, both of which have been implicated in the pathogenesis of PD by numerous studies [30, 33, 55]. Thus, our findings support the hypothesis that aberrant proteostasis contributes to the pathogenesis of PD.

In the healthy aged brain we identified a group of genes exhibiting inverse correlations between transcript and protein levels. Anticorrelation between transcript and protein readouts can be explained by the highly polarized cellular architecture of neurons, which allows spatial separation between mRNA and protein [38]. While some proteins are translated locally at their resident site, others are synthesized in the soma and transported along the axon/dendrites to their target location. This leads to a steady state in which the transcript resides in the soma, whereas most of the protein is either under transport in the axon or at the synapsis [38]. In these cases, since brain tissue samples vary in relative grey/white matter content and therefore also in relative soma/axonal content [39], readouts of transcript and protein levels will be anticorrelated across-samples. Specifically, samples enriched in somas will show a high relative transcript/protein ratio, whereas samples enriched in axons will show a low relative transcript/protein ratio. In line with this hypothesis, genes showing negative correlation between transcript and protein levels in healthy ageing were significantly enriched in synaptic vesicle related pathways. Synaptic vesicle proteins were indeed shown to be preferentially translated in the cell body and undergo axonal transport to the synapses [22, 29], consistent with a spatial compartmentalization of transcripts and their protein products. We also observed that the top negatively correlated genes in healthy ageing were highly positively correlated in the infants, which may reflect a more homogenous distribution of somata and axons and/or reduced axonal transport during development, likely due to immature neuronal morphology [21, 45].

Interestingly, genes showing inverse mRNA-protein correlation in PD were not significantly enriched in synaptic function compared to healthy ageing. At least two factors may contribute to this phenomenon. First, disruption of axonal transport has been shown to occur in the PD brain (see [52] for a review). This would decrease the spatial separation between transcript and protein, thereby blunting the negative correlation across samples. Second, the PD brain, including the prefrontal cortex, is characterized by neuronal and synaptic loss and a relative increase in glial populations [39]. It is therefore conceivable that, if the anticorrelation signal originates from neurons, it may be diluted as a result of these changes in cellular composition. Genes showing inverse mRNA-protein correlation in PD were enriched for subunits of the proteasomal complex compared to both infants and neurologically healthy aged individuals. This finding suggests that these proteins become specifically more polarized in PD, with accentuated spatial separation of transcript and protein between soma and axon. The ubiquitin-proteasome system has a crucial role in maintaining synaptic proteostasis and modulating neurotransmission and has been shown to be enriched at the synapses [12, 16, 24, 46, 48]. Moreover, studies in mice have shown that some proteasomal subunits are translated locally at the synapses, whereas others are translated in the soma and transported to the synapses [11, 22]. Furthermore, our data indicate that the spatial mRNA-protein separation is uneven across the proteasomal subunits, suggesting a potentially altered stoichiometry of the synaptic proteasome in PD neurons. The formation of an alternative proteasome complex consisting of an additional *α* − 4 subunit (*PSMA7*) in place of an *α* − 3 (*PSMA4*) has been shown to be involved in cellular adaption to environmental stress [40]. These subunits showed a marked disparity in their correlation values in the PD brain (*r*_*PSMA7*_ = —0.51; *r*_*PSMA4*_ =− 0.17).

Interestingly, the PD brain was characterized by increased positive mRNA-protein correlation for genes encoding components of the mitochondrial respiratory chain. We and others have shown that quantitative and functional respiratory chain deficiencies characterize the PD-brain, including the prefrontal cortex [18, 44]. It is possible that a tighter relationship between transcription and translation of at least some of the mitochondrial respiratory chain subunits allows for better regulation in a highly strained system lacking spare capacity. Positive correlations for widely expressed genes in neurons were also observed by [38].

The findings of this study should be interpreted in light of several limitations. First, post-mortem RNA degradation in our samples may partly contribute to low correlations between mRNA and protein. Proteins are generally more resilient to post-mortem degradation and survive for longer periods than RNA. However, since there is no reason to assume that RNA degradation would be systematically different between our groups, this factor is unlikely to confound our results of differential transcript-protein correlation between groups. Second, due to the lower sensitivity of proteomics, our dataset was constrained to only 1,400 proteins. Thus, our findings are not necessarily representative of the entire genome. Third, the sample size for the YG group (*N* = 4) was small due to the limited availability of this type of tissue, limiting the generalizability of the ageing-associated findings. Nevertheless, the infant group did recapitulate the previously observed high positive correlation for the vast majority of genes [54], suggesting the samples are representative for transcript-protein correlation in the infant brain.

In summary, we show that the PD brain is characterized by altered coupling between the transcriptome and proteome, compared to neurologically healthy ageing. This altered relationship between mRNA and protein levels is consistent with an extensive, possibly proteome-wide, impairment of proteostasis, and strongly supports the hypothesis that aberrant proteasomal function is implicated in the pathogenesis of PD. Moreover, these findings have important implications for the correct interpretation of transcriptomic studies in this field. Gene expression studies are extensively used to identify disease-related pathways in ageing and neurodegeneration, and it is generally assumed that observed differences in mRNA levels reflect differences at the protein level. If the relationship between transcript and protein is altered in PD, this should be accounted for when interpreting the molecular impact of differential gene expression in the patient brain.

## Data Availability

The datasets supporting the conclusions of this article are included within the article and its supplementary files.
The source code for the analyses and the protein and transcript counts can be requested from the corresponding author.

## Supporting Information

**S1 Figure.**
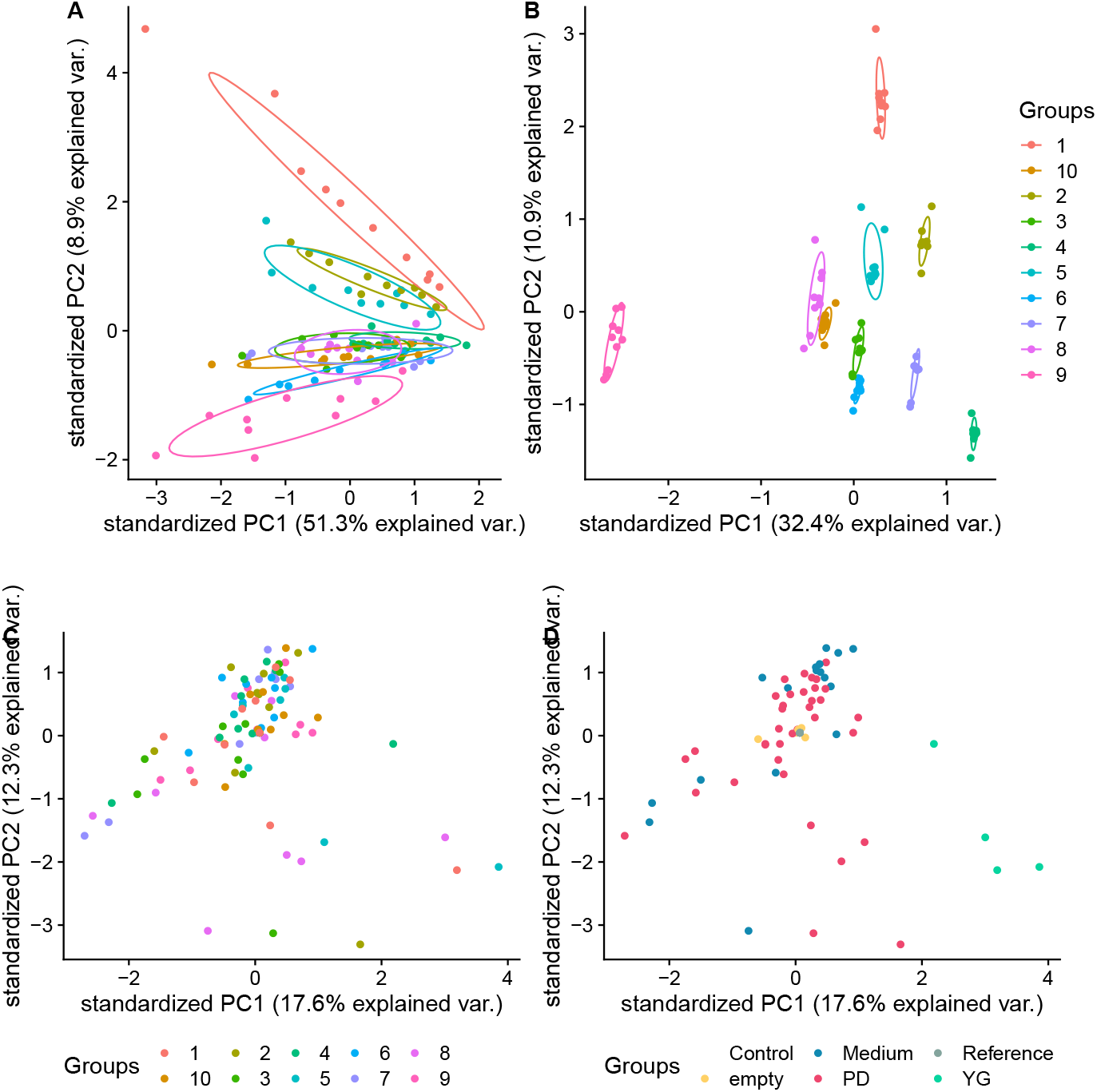
Principal component analysis visualized for all TMT samples. Data points represent samples spanned by the first (x-axis) and second (y-axis) component of principal component analysis on raw protein intensities (**A**), quantile normalized protein intensities (**B**) and scaled batch corrected protein intensities (**C** and **D**). Colouring indicates the TMT batch of the sample for A, B and C and the sample’s condition for D.

**S2 Figure.**
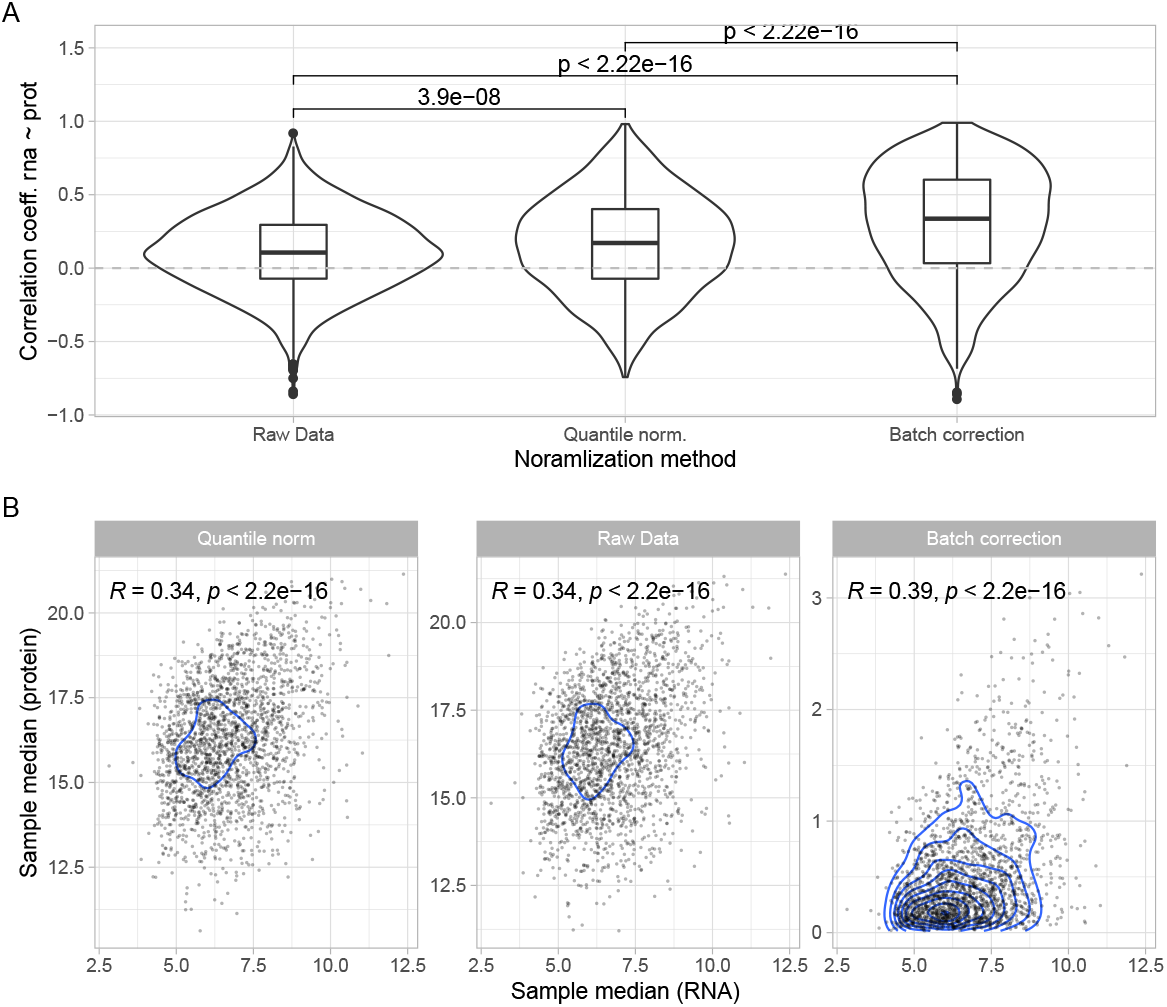
Batch correction of proteomics data improves correlation with RNA. **A:** Distribution of gene-wise correlation coefficients (RNA ∼ protein) (y-axis) are displayed for the three normalization approaches of protein intensities (x-axis). **B:** Comparison of correlation between sample-median RNA expression (x-axis) and sample-median protein expression (y-axis) for the three different protein intensity normalization approaches (facets).

**S3 Figure.**
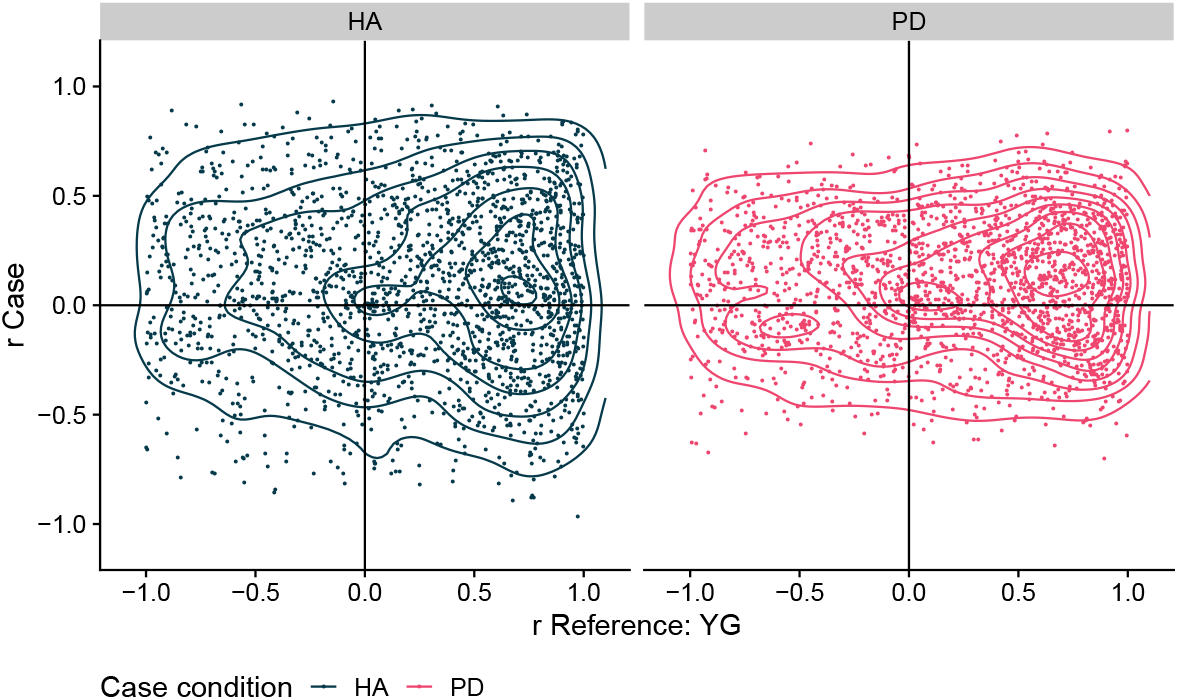
Distributions of RNA-protein correlations in a comparison between groups. Two-dimensional density plot displaying both distribution and relationship between the reference YG (x-axis) and the ageing groups (y-axis): YG vs HA (dark blue, first panel), and YG vs PD (pink, second panel).

**S1 File**

**Cohort demographic and experimental information**

**S2 File**

**Correlation coefficients and gene ranking**

Contained are gene-wise Pearson correlation coefficients for each group (YG, HA, PD) as well as scorings used to rank genes in the pathway enrichment analyses.

**S3 File**

**Significantly enriched GO terms**

Enriched go terms for each gene scoring are listed in respective sheets.

## Acknowledgments

Mass spectrometry-based proteomic analyses were performed by (The proteomics Unit at University of Bergen (PROBE)). This facility is a member of the National Network of Advanced Proteomics Infrastructure (NAPI), which is funded by the Research Council of Norway INFRASTRUKTUR-program (project number: 295910).

We are grateful to patients and their families for participating in our research. We would also like to thank our colleagues at the Neuromics group for the fruitful discussions.

## Author contributions

- Conceptualization: F. Dick, G. S. Nido, C. Tzoulis
- Methodology: F. Dick. G. S. Nido
- Software: F. Dick
- Formal Analysis: F. Dick, G. S. Nido
- Data Curation: F. Dick, G. S. Nido
- Original Draft Preparation: F. Dick
- Review and Editing: F. Dick, C. Tzoulis, G. S. Nido
- Visualization: F. Dick
- Provision of material: G. W. Alves, O. Tysnes
- Supervision: C. Tzoulis
- Project Administration: C. Tzoulis
- Funding Acquisition: C. Tzoulis

## Data availability

The datasets supporting the conclusions of this article are included within the article and its supplementary files. The source code for the analyses can be requested from the corresponding author.

## Funding

This work is supported by grants from The Research Council of Norway (288164, ES633272) (https://www.forskningsradet.no/en/) and Bergen Research Foundation (BFS2017REK05) (https://mohnfoundation.no/engelsk-rekruttering/?lang=en). Both of these were received by CT. The funders had no role in study design, data collection and analysis, decision to publish, or preparation of the manuscript.

## Competing interests

The authors have declared that no competing interests exist.

## Notes

### Competing Interest Statement

The authors have declared no competing interest.

### Author Declarations

Regional Ethics committee of Western Norway (REK 2017/2082, 2010/1700, 131.04)

